# The experiences of general practitioners regarding communication with patients from different cultural backgrounds and/or low socio-economic status: a qualitative interview study

**DOI:** 10.1101/2025.03.27.25322990

**Authors:** Dionne N. Zwinkels, Hannah A. Bijl, Reinier C.A. van Linschoten

## Abstract

**Introduction:** Effective communication between general practitioners (GPs) and patients is essential for high-quality healthcare, improved patient outcomes and physician satisfaction. However communication barriers related to cultural differences and low socio-economic status (SES) can negatively impact patient care. This study explores the experiences of GPs in navigating communication challenges with patients from diverse cultural and socio-economic backgrounds, aiming to identify strategies to improve these interactions.

**Methods:** This study used a qualitative phenomenological approach to explore GPs’ experiences with communication challenges with patients from diverse cultural backgrounds and/or low socioeconomic status. Semi-structured in-depth interviews were conducted with GPs from Jans Huisartsen, a general practice in Rotterdam. Data were analyzed with thematic analysis.

**Results:** A total of 13 GPs from Jans Huisartsen were interviewed. Key challenges included language barriers, different expectations, and different cultural norms. Consequences of communication challenges included misunderstanding, lower medication adherence, less shared decision-making, and decreased patient satisfaction. To improve communication, GPs employed strategies such as simplifying language, using translators, and personalizing consultations.

**Conclusion:** This study highlights the experiences of GPs regarding how cultural diversity and SES affect effective communication. The results underline the importance of addressing specific challenges faced by these patients, particularly by fostering shared decision making and re-evaluating the role of family translators. Enhancing GP training on communication with culturally diverse patients or patients with low SES and promoting the use of professional translators could improve patient understanding, adherence and satisfaction, ultimately contributing to more equitable and effective healthcare for these vulnerable populations.

## Introduction

Effective communication between doctors and patients is essential for high-quality healthcare, showing benefits for both patients and general practitioners (GPs). Research indicates that good communication leads to higher patient satisfaction, which in turn increases physician satisfaction and reduces work-related issues such as burnout and stress.(1, 2) Moreover, a strong doctor-patient relationship is associated with better physiological, behavioral, and subjective patient health.(3) Effective communication enhances the exchange of information and adherence to treatment, ensuring that consultations are productive and meet patient needs. This is especially important in general practice where long-term relationships with patients are built.(4)

The purpose of doctor-patient communication is to build a good relationship, exchange information, and make treatment-related decisions. Good communication can be defined as a process that effectively combines informative (cure-oriented), affective (care-oriented) components, along with patient-centeredness.(5) Informative communication involves sharing and seeking medical information to ensure patient understanding and is crucial for accurate diagnosis and treatment planning. Affective communication focuses on empathy, treating patients as individuals rather than cases, fulfilling their need to feel known and understood. Patient-centeredness ties these elements together by ensuring that care align with the patient’s needs, preferences, and values. All three components are necessary for effective doctor-patient communication.(5, 6)

Cultural differences and low socio-economic status (SES) can impede effective communication between doctors and patients and negatively affect the quality of care.(7, 8) Communication barriers related to cultural or ethnic backgrounds are differences in explanations of health and illness, cultural norms and values, preferences in doctor-patient relationships, such as whether a personal relationship with the GP is valued or whether a more hierarchical, professional distance is preferred, biases from both doctors and patients, and language barriers.(7) These issues can lead to less affective and informative communication between doctors with patients from ethnic minorities, undermining the effectiveness of care.(7) For instance a study demonstrated that non-Western patient groups are less participative during medical consultations than Dutch patients, while participation is essential for understanding and adherence. The communication skills of the GP had the highest impact on participation and patient satisfaction.(9) Further research indicates that intercultural communication often leads to misunderstandings and lower quality of care, which negatively affects the satisfaction and adherence of patients with a migration background.(10-15)

SES is another important factor influencing doctor-patient communication. SES refers to an individual’s social and economic position within society, typically defined by factors such as income, education level, and occupation status. People with lower SES tend to have lower levels of education and work in lower-skilled jobs, which often correlates with lower incomes. These combined elements influence both their financial position and their social status.(16) Individuals with lower SES generally have poorer health and a shorter life expectancy than those with higher SES.(17-19) These health disparities can shape patients’ perceptions and expectations of healthcare. Research shows that people with lower SES more often have negative experiences with healthcare services and are less satisfied with the care provided.(9) SES also impacts doctor-patient communication. Patients with lower SES more frequently experience communication problems such as misunderstandings during medical consultations compared to those with higher SES. This is primarily due to lower health literacy, reduced trust, and different expectations.(20) These problems can lead to lower satisfaction with health care and lower adherence.(8, 9)

Most research on communication in healthcare has been done from the perspective of patients. To our knowledge there are no studies from the perspective of doctors regarding communication with patients with low SES and/or diverse cultural backgrounds. The aim of this research was to explore GPs’ experiences with communicating with patients from diverse cultural backgrounds and/or lower SES. By understanding these experiences, targeted strategies can be developed to enhance communication and improve the quality of care.

## Methods

### Qualitative approach and research paradigm

This study used semi-structured in-depth interviews and thematic analysis to explore the experiences of GPs with communication with patients from different cultural backgrounds and/or low SES. The research approach in this study was phenomenological, focusing on understanding the lived experiences of GPs. This approach acknowledges the existence of multiple subjective realities, as each participant’s experience and interpretation are unique.

### Researcher characteristics and reflexivity

The researcher of this study was a medical student from Erasmus University who started her study in 2018. This research was part of the Master of Medicine. The researcher is Dutch with a Western cultural background, residing in Rotterdam. The interviewer had limited experience with conducting interviews. She knew some of the participants, as she worked as a doctor’s assistant in the practice where the study was conducted. Familiarity with some participants, due to prior work as a doctor’s assistant, might affect objectivity in data collection and interpretation. The researcher’s cultural perspective could shape question framing and response interpretation.

### Context

We conducted the study with GPs from Jans Huisartsen. This is a general practice organization with three practices in Rotterdam. Rotterdam is a multicultural city with a diverse population, characterized by a wide range of cultural and ethnic backgrounds. The neighborhoods of the three practices vary considerably in terms of SES, ethnic and cultural backgrounds of the residents. In the neighborhood in the south of Rotterdam, the proportion of residents with a non-Western migration background is 57%, compared to 39% and 38% in the city center and the eastern neighborhood, respectively. Considering education levels and household income, 39% of residents in the south have no diploma, and 51% have a low income. In the center of Rotterdam, these figures are 18% and 40%, and in the east, they are 24% and 51%.(21)

### Sampling strategy

The study population consisted of GPs working at Jans Huisartsen. GPs could work at one or more of the locations. All the GPs at Jans Huisartsen were invited to participate in the study via email. There were no exclusion criteria. We aimed to include all eligible GPs in our study. The sampling process took place in July and August 2024.

### Ethical issues pertaining to human subjects

This study was conducted by the Department of General Practice of the Erasmus MC in collaboration with Jans Huisartsen. A waiver for the Medical Research Involving Human Subjects Act was obtained from the non-WMO review committee of the Erasmus MC. Participants were informed about the study’s purpose, the voluntary nature of their participation, and their right to withdraw at any time without consequences. All GPs who agreed to take part in the interviews signed and returned their written consent before the start of the interview. Participant confidentiality was ensured by pseudonymizing the data and securely storing interview recordings and transcripts.

### Data collection

Data was collected through in-depth semi-structured interviews conducted by the first author. A topic guide was used for the interviews to gain insight into the doctor’s perspective regarding the communication with patients with lower SES and/or other cultural background, the challenges they encounter and the strategies they employ to address these challenges (Supplementary File 1). The topic guide was used for all interviews; however, it was adjusted as more interviews were conducted to explore emerging themes in greater depth. The researchers used prompting and probing techniques, such as asking open-ended questions or requesting participants to elaborate on specific points, to delve deeper into topics mentioned during the interviews. The interviews took place in a consultation room at the practice where the interviewed doctor worked that day. All interviews lasted around 20-30 minutes. The interviews were recorded with Microsoft Teams.

### Data processing & analysis

Interviews were recorded and transcribed verbatim and entered into MAXQDA. The transcript of each interview was pseudonymized, and each participant was given a code number. A coding tree was developed by the researcher and reviewed and refined by the research team. The analysis began with open coding to identify key statements and phrases pertinent to the research questions. This was followed by axial coding to connect these codes to broader (sub)themes. This process involved identifying patterns and themes related to the experiences of GPs with communication challenges involving patients from different cultural backgrounds and/or low SES.

### Techniques to enhance trustworthiness

To ensure the trustworthiness of this study, peer reviewing was employed. The first author initially conducted the coding independently, after which another researcher coded a subset of transcripts. Based on this process, the coding tree was further refined. Subsequently, the first author recoded all transcripts using the updated coding tree.

## Results

### Characteristics of participants

The study included a total of 13 GPs, of whom 7 (53.8%) were female and had an average age of 38.2 years (Table 1). Most participants were around 30 years old, with one outlier being a GP aged 69 years. Most participants (69.2%) were of Western ethnicity, followed by Asian (23.1%) and Arabic (7.7%). All grew up in the Netherlands and indicated they had a Western cultural background. The participants had an average of 7.2 years of professional experience, a mean of 1.1 years at Jans Huisartsen. The older GP had 41 years of experience as a GP and had been working at Jans Huisartsen over 3 years. Nearly all participants (84.6%) had experience with populations with low and high SES.

**Table 1.** Baseline Characteristics.

| **N= 13** | **n (%) / years (SD)** |
| --- | --- |
| Female | 7 (53,8) |
| Mean age | 38,2 (9,4) |
| Ethnicity |  |
| *Western* | 9 (69,2) |
| *Asian* | 3 (23,1) |
| *Arabic* | 1 (7,7) |
| Cultural Background |  |
| *Dutch (Western)* | 13 (100) |
| Mean work experience as GP | 7,2 (10,2) |
| Mean work experience at Jans | 1,1 (0,8) |
| Experience low SES* | 12 (92,3) |
| Experience high SES* | 12 (92,3) |
| Experience both low and high SES* | 11 (84,6) |

\*Working or have worked as a GP in a low or high SES neighborhood

### Main findings

The main finding of this study was that most of the interviewed GPs thought that a patient’s low SES or different cultural background had a considerable influence on doctor-patient communication. Data analysis revealed three main categories: challenges in communication, consequences of communication challenges, and improving communication. The categories were divided into various themes and subthemes (Figure 1). An overview of the codes used with definitions and example quotes can be found in Supplementary File 2.

**Figure 1.**
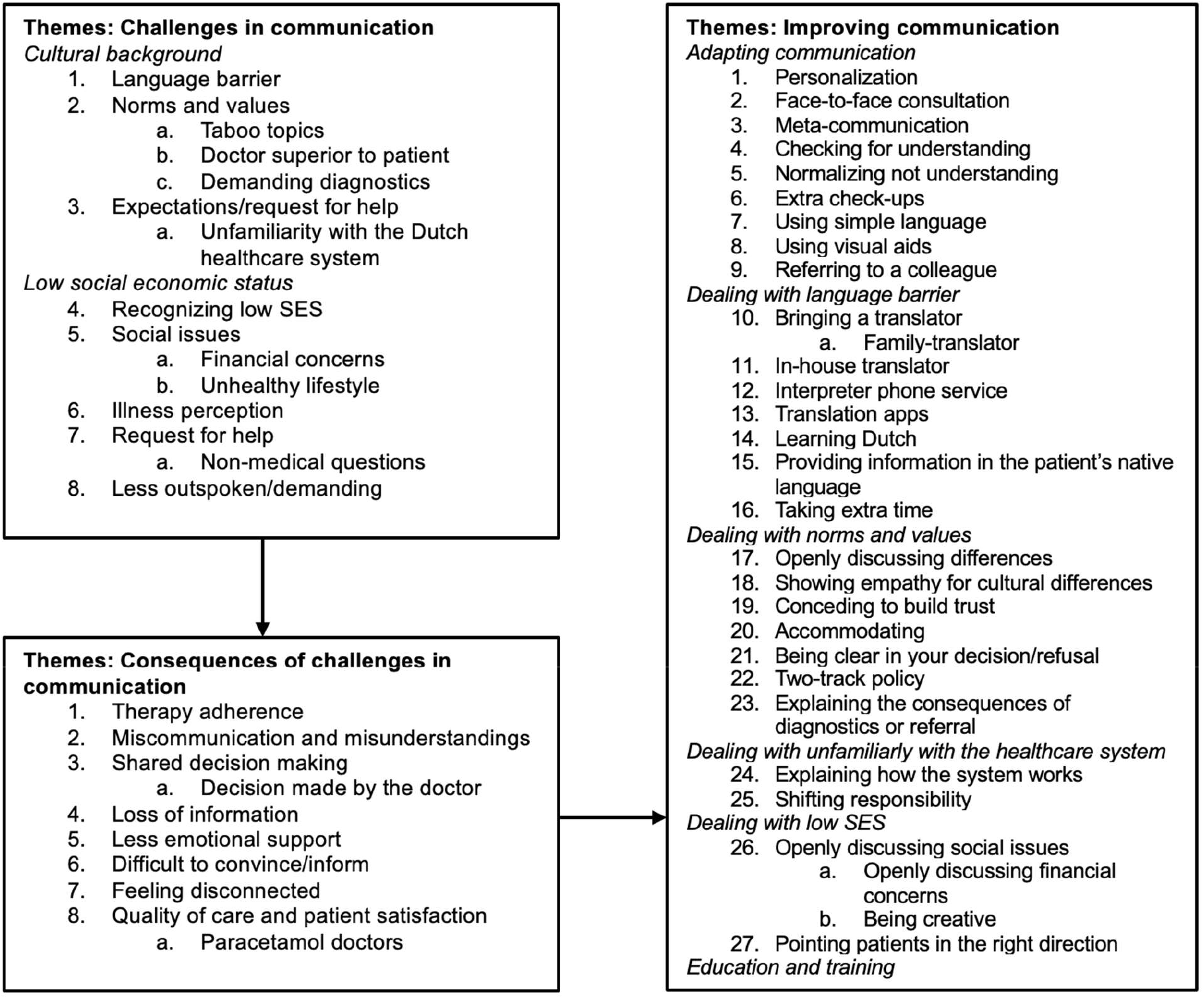
Themes and subthemes.

### Challenges in communication

All participants identified challenges in communication with patients of low SES or different cultural backgrounds.

For patients with a different cultural background, GPs highlighted language barriers, differing norms and values, and mismatched expectations as major obstacles. Language barriers complicated understanding the patient’s concerns, with GPs noting that translation apps lose nuance and interpreters may not always provide accurate translations. One GP stated: *“It’s harder to pinpoint the core issue*… *It’s more difficult. You can easily put in a stitch, but it’s the parts that are a bit more grey that are more challenging*.*”* These issues often lead to increased reliance on diagnostics: *“So, you just end up listening, watching, looking, and measuring because you can’t get the information out of them*.*”*

Cultural differences also surfaced in discussions on sensitive topics like mental health or contraception, with patients holding different views. For example, a GP stated: ‘*‘Sexual problems are often difficult to discuss, that can also be a taboo. But for some complaints it is important to discuss, so then you have to’’* and another GP: *‘‘I think especially psychological things are less talked about, especially feelings and emotions’’*. Moreover, GPs thought that patients from different cultural backgrounds were sometimes used to viewing the doctor as superior to the patient.

Additionally, GPs mentioned that patients from other backgrounds may have expected quicker referrals or treatments: ‘*‘They are demanding, so they want to have certain examinations, they want to have certain medications. Because they are used to…the fact that in their own country, yes then they pay for the doctor and then they are allowed to go to the specialist. And then they also just get the examinations they ask for*’’. They are unfamiliar with the Dutch healthcare system, particularly in terms of insurance: ‘*‘And what a lot of people don’t understand is the socialist healthcare system*…*People come up with the idea ‘I pay premiums, so I’m insured’’. That’s a very common statement*’’, which could lead to prolonged explanations and discussions as mentioned by some GPs.

GPs reported using various methods to identify or assess whether a patient has a low SES. Some mentioned that they ask directly about a patient’s SES, either during an introductory consultation or when it seems relevant to the complaint the patient presents. Others do not explicitly ask, but estimate a patient’s SES based on their vocabulary and manner of speaking. For patients with low SES, GPs noticed that these patients typically presented more physical complaints and struggled to articulate their concerns. Social issues, particularly financial problems, further complicated care, as patients may have been unable to afford recommended treatments: *“And often, what you run into are the costs*… *and then they say, ‘Well, I actually can’t afford that*.*”* Stress related to financial difficulties often manifests as physical symptoms. However, GPs also observed that patients with low SES often had a lower illness perception. For example, they might not recognize the connection between their emotional or financial stress and the physical symptoms they experience: *‘‘They don’t quite understand that the stress they have can also make them feel sick*.*’’* GPs also noted that the health of patients with low SES was generally worse due to lifestyle factors like alcohol use and hypertension. Due to these factors patients with low SES visited the GP more frequently.

Some GPs mentioned that the concerns of patients with low SES were often non-medical but logistical questions, for example related to housing, parenting, and finances. This can make it challenging to determine what can be done for such a patient and where the GP’s responsibility lies: ‘*‘So there are really more logistical challenges. I do sometimes find it difficult to determine to what extent I am responsible for that’’*. Last, some GPs thought that they were more easily reassured than high-SES patients and that these patients were less outspoken and demanding compared to high-SES patients.

### Consequences of communication challenges

GPs stated that the communication challenges can have consequences for the GP, but more importantly the patient. Some GPs mentioned that difficult communication led to lower medication adherence.

Reasons for this include patients not understanding the importance of the medication, especially when managing cardiovascular risk, because people have difficulty taking medication without having immediate symptoms. Other reasons for non-adherence includes due to side effects or not understanding the medication instructions. This issue was believed to be particularly prevalent among patients with low SES. Miscommunication and misunderstandings often stemmed from language barriers or lower cognitive abilities, which could lead to dangerous health outcomes. One GP noted: *“Yes, it sometimes surprises me as well. It’s usually not major things, but when I agree on something with someone, and then they completely misunderstand it or come back with something entirely different from what we had agreed upon*.” And another GP: “*A specific example is taking the contraceptive pill after having sex*.”

Shared decision-making (SDM) is important in general practice, but the GPs in this study had differing views on how SES and cultural backgrounds affect this process. Some male GPs reported applying SDM with all patients, regardless of background, and reported having no problems with this. Others found it more difficult due to language barriers or because patients did not understand that they had a choice, as they assumed the doctor held all the knowledge. In these cases, GPs took a more authoritarian approach, making decisions on behalf of the patient: “*With patients from a low SES background, you tend to notice more quickly, like, ‘Well, you’re the doctor after all*.*’ So, you adopt a bit more of an authoritative stance, like, ‘I think this is good for you, so I’m going to make this referral or prescribe this medication*.”

Doctors also noted that medical information was lost due to language barriers or lower cognitive abilities. Moreover, the presence of an interpreter, as a solution for a language barrier, could sometimes lead to more communication directed toward the translator instead of the patient. Additionally, a language barrier could make it more challenging for GPs to provide emotional support: ‘*‘When someone has a language barrier it does make it more difficult I think. You can’t deal with emotions well’’*.

Another consequence of difficult communication is that patients are less easily convinced of their illness narrative or the importance of treatment: “*No matter how simply you explain it to them, in my opinion, I can’t explain it any more clearly, and yet you see that they still keep coming back or calling*.” Furthermore, patients may feel disconnected from society due to its complexity, which presented challenges in explaining societal aspects of healthcare. For instance, using a patient portal could be very complex for individuals with lower educational backgrounds.

Last, communication challenges may have implications for the quality of care and patient satisfaction. Participants had differing opinions on the magnitude of this impact. Some GPs felt that patients from different cultural backgrounds or lower SES might be less satisfied with the care they receive due to differing expectations or miscommunication. However, GPs often did not notice this directly, as dissatisfaction was often reported later to the assistant, or the patient simply didn’t return. One GP believed that the quality of care for this patient population is indeed lower: *“I do think that the quality of care they receive is lower. Also because we don’t have the patience or the time to explain everything properly*.’’ When patients were dissatisfied, it was often because they did not receive what they expected from the GP, such as a referral to a specialist or an MRI. Some patients even refer to Dutch GPs as “*the paracetamol doctor*”, as the Dutch GP did nothing else than prescribing paracetamol. On the other hand, some GPs believed that this patient group can be particularly grateful, loyal, and satisfied, as long as the GP takes the time to engage with them. A GP stated: ‘*‘I just found that there is a lot of gratitude and a lot of satisfaction, but that was also, I worked there 4 days a week so then you were also just really the point of contact and I took the time for those people. I think they need that a lot*’’.

### Improving communication

Several common strategies were identified by nearly all interviewed GPs for improving communication with patients with low SES or from different cultural backgrounds. They aimed to personalize consultations by sharing examples from their own lives, by telling about their own cultural background for example, opting for face-to-face consultations rather than digital consultations, employing meta- communication, checking for understanding by asking patients to explain what they have understood, normalizing situations where patients did not understand, planning extra check-ups, focusing on the use of simple language with short sentences and minimal jargon, using visual aids to clarify information, providing written information at the end of the consultation for patients or their family members to review at home, and referring patients to a colleague when communication becomes particularly challenging.

GPs highlighted specific strategies for addressing language barriers. They often encouraged patients to bring a translator, typically a family member, to consultations. When this was not feasible, a practice staff member was sometimes enlisted as a translator, or translation services and apps were used. GPs frequently expressed difficulty in determining their responsibilities in these situations: ‘‘*I sometimes find it difficult to determine to what extent I’m responsible for that”* and *“Honestly, a patient who comes to the doctor wants to be helped. So if their Dutch isn’t good, they should arrange for a translator*.’’ Additionally, two GPs suggested that patients should simply learn Dutch: ‘*‘I think it’s also partly their own responsibility. You moved to a country where you don’t speak the language and you don’t speak English. If you want to get good care then you have to make sure that you make it understandable for yourself*’’. Some GPs mentioned that they often provided information in the patient’s native language and emphasized the importance of taking time with patients experiencing language barriers when possible.

GPs also mentioned various strategies for addressing differences in norms and values. First and foremost, they deemed it important to openly discuss differences and demonstrate empathy for cultural backgrounds by attempting to understand the patient’s situation: ‘‘*What often helps me is to put myself in their shoes, even just a little. They aren’t used to anything else. If you look at it from their perspective, you know that they are used to going to a specialist quickly and getting all the tests; that’s normal for them. And when they come here and a doctor says no, that’s not allowed, you just have to wait and do nothing, you have to be a bit considerate of that, at least*.’’ Patients with certain expectations or specific requests that did not align with those of the GP could generally be expected to be involved in SDM, although their wishes may not always have been fulfilled. Two GPs noted that this can depend on the day: ‘‘*Of course, there are days when you think you couldn’t care less. If you really want that referral, I don’t feel like arguing about it’’*. Some GPs mentioned that they also sometimes conceded to gain the patient’s trust. However, overall, GPs noted that they tend to be strict regarding expectations for procedures like MRIs or referrals and try to find a middle ground with patients through informative discussions. When GPs did not agree with a patient’s requests, they mentioned that they find it crucial to communicate this as transparently and clearly as possible. Some GPs adopted a two-track policy, where they managed the patient’s expectations by proceeding with the medically indicated course of action while simultaneously ordering additional tests or referrals to satisfy the patient’s desire for reassurance. Others attempted to explain the implications of unnecessary further investigations or referrals to patients. Some GPs mentioned that explaining could help, while others indicated that this often needed to be repeated at each consultation, as patients struggled to fully understand or retain the information.

Regarding unfamiliarity with the Dutch healthcare system, GPs emphasized that providing clear explanations and investing time in patient education is the most effective approach. Additionally, they found it helpful to shift responsibility by clarifying that they, as GPs, were not accountable for how the Dutch healthcare system operates: ‘‘*But I also sometimes say that I am a caregiver and that this is my job, but that I don’t have control over the healthcare system. That can also help. It’s like using a sort of lightning rod*.’’

Specifically for patients with lower SES, it was essential for GPs to openly discuss social and, more importantly, financial issues. It required reativity from GPs to collaboratively find solutions for problems that may have been beyond their control. ‘’*With a few euros, you can buy a bag of potatoes and a box of frozen frikandellen. Yes, I can tell someone to eat healthier, but that’s just not possible. It’s really about accepting that different rules apply here. So I think you have to be a bit more creative*’’. For this patient population, GPs also mentioned that it was important to guide patients toward appropriate resources

The above strategies employed by GPs to improve communication were mainly gained from work experience. GPs mentioned that there was limited focus on this topic during their residency, but that from experience they learned how to communicate with these patients: ‘*’GP residency is only 3 years and there’s a lot to go into that. It’s also what you learn in your own population of course. Gradually you get to know your people a little bit. That’s where you kind of figure it out*’’,

Two broad approaches to dealing with communication challenges emerged among the GPs interviewed. One group emphasized patient responsibility, encouraging patients to learn Dutch, arrange their own translators or accept the structure of the Dutch healthcare system. These GPs saw their role as primarily focused on providing medical care, occasionally shifting responsibility for communication challenges back to the patient or delegating cases to colleagues when barriers were too great. This group consisted mainly of male GPs, with one exception. The other group, mainly female GPs, prioritized a more patient-centred approach and sought to adapt their communication strategies by simplifying language, taking extra time or addressing underlying social factors. This group generally put more effort into bridging the gap between the patient and the healthcare system.

## Discussion

### Main findings

GPs identified several communication barriers when working with patients from diverse cultural or low SES backgrounds. Common challenges included language barriers, varying norms and values and unmet expectation. These challenges often led to miscommunication, reduced treatment adherence, and less frequent SDM, which negatively impacted both the informative and affective communication, ultimately affecting the quality of care and patient satisfaction. In response to these challenges, GPs reported a wide range of strategies to improve communication, such as simplifying medical language, personalizing consultations, and using techniques like ‘checking for understanding’. Additionally, some GPs noted the value of involving translators, though there were limitations to this approach, especially when family members or children were used as translators. In addressing communication challenges, there were two groups with a possible influence of gender. Female GPs were more likely to have a patient-centred approach, whereas male GPs were more likely to describe putting more responsibility on the patient. The focus on personalized communication and empathy, as well as the use of culturally sensitive approaches, was emphasized as important for improving patient-GP relationship, improving patient engagement and ultimately, the quality of care.

### Comparison with existing literature

The challenges identified by GPs in this study in communicating with patients from different cultural backgrounds align with the existing literature, which predominantly focused on patients’ perceptions regarding doctor-patient communication with diverse cultural backgrounds and low SES.(7, 8) Earlier research comparing the communication styles used by white physicians when interacting with white patients or ethnic minority patients revealed similar obstacles during cross-cultural consultations, including language barriers, differing cultural norms and values, and varying perceptions of illness and healthcare.(6, 7) A challenge identified by the GPs in this study is the unfamiliarity that many patients from different backgrounds have with the Dutch healthcare system. While this could fall under differences in norms and values, it is not explicitly described in the existing literature.

Patients with lower SES frequently face social issues, such as financial difficulties and unhealthy lifestyles, as noted by GPs in this study. These patients were observed to have a different illness perception and were generally less outspoken and demanding. Some GPs in our study indicated that this made communication with patients with low SES feel easier compared to patients with higher SES, as these patients tended to ask fewer questions and sought less detailed information. However, this perceived ease may come at a cost, as patients with low SES might receive less information and emotional support, potentially leading to feelings of disconnection.(22)

This observations aligns with prior research, which shows that patients with lower SES often receive less socio-emotional support, experience a more directive and less participatory consultation style, and are less involved in treatment decisions. These consultations frequently involve a higher percentage of biomedical discourse, reduced patient control over communication, and limited diagnostic and treatment information.(23) Furthermore, an earlier study found that patients with higher SES tend to ask more questions and seek more detailed medical explanations.(24)

Such challenges raise concerns about whether care for patients with lower SES aligns with the core values of general practice as outlined by the NHG (Dutch College of General Practitioners) and LHV (Dutch General Practitioners Association): person-centered, medical-generalistic, continuous, and collaborative.(25)

A key component of person-centered and collaborative care is SDM, which is widely recognized as essential in healthcare, enabling patients to participate in decisions about their health and treatment.(26) However, several GPs in our study acknowledged that they applied SDM less frequently patients with lower SES and those from diverse cultural backgrounds. This reluctance often stems from patients expressing a preference for the GP to make decisions, based on an assumption that the GP possesses superior knowledge. Additionally, GPs reported that they sometimes bypass SDM if they perceive a lack of illness perception among these patients or if language barriers make SDM challenging.

Research indicates that patient characteristics, such as self-efficacy, self-awareness, and illness severity, also impact engagement in SDM.(27) Patients may require support in areas like emotional management and communication to fully participate in decision-making. Barriers to SDM, particularly intercultural differences, language barriers, differing values regarding health, dissimilar role expectations, and biases, align with the results of our study and impact the quality of interaction between physicians and patients.(27)

Furthermore, patient-centered medicine advocates for self-determination and informed decision-making, emphasizing that involving patients in SDM promotes adherence to treatment and leads to better health outcomes.(28) This raises ethical questions regarding the justification for selectively applying SDM based on patients’ SES or backgrounds. When SDM is not consistently implemented, patients with culturally diverse backgrounds or lower SES may not fully experience the person-centered and collaborative care that general practice aspires to provide.

### Implications for research and practice

The findings from this study highlight several important considerations for both research and clinical practice. One of the key issues raised is the distinction between formal GP training and learning on the job. As many of the GPs in this study relied on practical experience to navigate communication challenges, it raises the question of whether formal training programs could be enhanced to better prepare practitioners for consultations with patients with low SES or different cultural backgrounds. Programs such as those offered by Pharos, which focus on culturally sensitive communication, could serve as a valuable resource.(29) Pharos provides training that helps healthcare professionals communicate more effectively with patients who have low literacy, a migration background, or refugee status, tailoring care to patients’ language, education, health literacy, and cultural background. The program focuses on understanding cultural differences in health beliefs, the role of the patient, caregiver, and healthcare provider, and the stressors that come with migration or low SES. It emphasizes the importance of open, effective communication, especially in motivating patients towards health-promoting behaviors and treatment adherence and helping patients with low health literacy.Training programs like these could play a role in bridging the gap identified in this research. Future research could assess the effectiveness of these existing training programs and explore ways to further optimize them to meet the specific needs of underserved patient populations.

Another important question is whether the communication strategies GPs use are sufficiently adapted to meet the needs of patients, particularly in cross-cultural consultations. The focus of this paragraph is to examine the various communication strategies used by GPs when interacting with patients from low SES or cultural backgrounds, assessing their effectiveness, potential pitfalls, and how these practices align with established guidelines for accessible communication.

As shown in Figure 1, many adaptations were made in communication with patients with low SES or culturally diverse patients. Guidelines on accessible communication for patients emphasize several strategies for improving comprehension, many of which align with the adaptations described by GPs in this study.(30) These guidelines recommend using clear and explicit language, such as speaking in short sentences and conveying only one message per sentence. Additionally, it suggests speaking slowly and clearly, allowing for sufficient pauses, using the patient’s own words where possible, and avoiding medical jargon and abstract terms for example. Repeating key information several times is also advised. These recommended approaches closely reflect the communication adjustments reported by GPs in this study, who sought to make consultations more understandable and effective for patients with low SES and culturally diverse patients.

One method frequently used by GPs is “checking for understanding,” in which the doctor asks the patient to explain in their own words what they have understood. Research has demonstrated that this method is effective and leads to better patient health outcomes.(31) It is recommended that this technique is not only applied at the end of the consultation but also throughout the discussion to ensure comprehension.(30)

Earlier research found that doctors try to simplify their language with patients with lower SES.(20) This matches the results of this study, as several doctors indicated that they try to explain medical information to patients with low SES by simplifying their language use and using illustrations. However, while illustrations can aid in comprehension, not all images are effective, especially for patients with low literacy levels. Studies have shown that individuals with lower health literacy tend to process images differently from highly literate individuals, due to differences in abstract reasoning, spatial awareness, and cultural background. In fact, simple drawings made by doctors are often not well understood by patients.(32) Therefore, using carefully designed, culturally sensitive illustrations is crucial for ensuring that visual aids effectively support the communication of medical instructions.

In addition to simplifying language, GPs in this study also described making an active effort to show greater empathy toward patients with low SES and cultural diverse patients by trying to place themselves in the patient’s situation. It is well-documented that patients who perceive their doctor as empathetic report higher levels of satisfaction.(33) Building a trusting relationship between the GP and patient has been shown to positively influence health outcomes.(34) Therefore, fostering empathy and trust with patients from all SES backgrounds is crucial for improving the effectiveness of the care provided.

The use of family members as translators was frequently mentioned as a practical solution to language barriers by the GPs in this study. While it is commonly used, the practice is not without drawbacks. GPs in this study acknowledged that at times, they end up focusing more on the translator than on the patient, which can reduce the attention given to the patient’s needs and concerns. Moreover, they noted that relying on family translators, especially young children, can lead to issues such as inaccuracies, confidentiality breaches, and emotional strain, which affect the quality of the information relayed during medical consultations. This is supported by research showing that young informal translators often experience negative outcomes, such as feeling burdened by the responsibility, encountering technical language barriers, and feeling discomfort when discussing sensitive or taboo topics.(35) Young adult informal interpreters reported three times as many negative experiences as positive ones when reflecting on their role as child interpreters. They noted how, as children, they often left out information, translated inconsistently, and lacked the emotional maturity to handle the responsibility. These findings suggest that healthcare providers and policymakers should be cautious about using family members, particularly children, as interpreters and emphasizes the importance of formal interpreters.

Time constraints in general practice consultations, particularly with patients of diverse cultural backgrounds, are known to be higher, as recent studies have shown that the workload for GPs is generally greater for non-native patients compared to native Dutch patients of similar age, gender, and SES.(36) GPs often feel pressured to condense complex medical information into short consultations, leaving little room for ensuring patient understanding. However, research has shown that longer consultations do not necessarily result in greater patient satisfaction, as satisfaction is influenced more by the quality of interaction than by consultation length alone.(37) This was confirmed by several GPs in this study, who did not indicate that more time would necessarily make consultations easier. Future research could explore how consultation strategies, rather than duration, might be adapted to better support patient understanding and lead to improved outcomes.

Finally, the issue of whether patients should take responsibility for learning the Dutch language was raised. Encouraging language learning could foster greater resilience and integration into the healthcare system, empowering patients to engage more fully in their care. However, t his raises ethical questions about the extent to which GPs can or should expect patients to shoulder this responsibility. Exploring how language acquisition programs and resources can be made accessible to underserved populations, and the role of healthcare providers in supporting this, could be valuable avenues for future research.

### Strengths and limitations

By using a phenomenological approach and semi-structured interviews, the study provided an in-depth exploration of GPs’ lived experiences. This design allowed participants to share their personal and professional challenges in communicating with patients, providing rich and nuanced insights. The qualitative nature of the study was well-suited to capturing GPs’ perceptions and experiences, which may not be fully captured through quantitative methods.

Furthermore, the results offer practical insights into the strategies GPs use, such as simplifying language and personalizing consultations. While it is not possible to determine from this study whether these approaches are definitively effective, they highlight common methods that healthcare professionals facing similar challenges in communicating with patients from low SES or culturally diverse backgrounds may consider implementing. Thereby we critically evaluated the effectiveness of these strategies and the study suggests improvements for practice, examining what GPs could do more or less of to enhance communication with patients from low SES or culturally diverse backgrounds. These real-world clinical experiences provide valuable guidance for GPs looking to enhance their communication skills, improve patient engagement, and ultimately raise the quality of care. Lastly, the study underscores the potential for formal training to support GPs in refining their communication strategies, indicating opportunities for further professional development and systemic improvements in primary care.

Despite these strengths, there are several limitations to this study. The sample size was relatively small, with all participants recruited from a single practice in Rotterdam, which may limit the generalizability of the findings. As a result, there is a risk of selection bias, and the results are likely specific to the context of Jans Huisartsen, rather than being applicable to GPs in other practices or regions. Furthermore, all the participating GPs identified as having a Western cultural background and were raised in the Netherlands, resulting in limited diversity in perspectives. Additionally, the Western perspective of the researcher may have influenced the research process. This potential bias could shape how the researcher interpreted the experiences of both the GPs and their patients, particularly when it comes to cross-cultural interactions. The researcher may have been less attuned to the subtleties of non-Western communication norms, potentially leading to an incomplete understanding of the communication barriers described. It is important to acknowledge this positionality as it may have affected both the framing of interview questions and the analysis of the results. Although familiarity with participants was initially considered a potential limitation, it did not prove to be an issue, as only one GP was personally known to the researcher.

### Conclusion

Overall, the findings underscore the importance of understanding and addressing the unique needs of patients from diverse cultural backgrounds and low SES, advocating for improved training and resources for GPs to facilitate effective communication and, ultimately, better health outcomes for these vulnerable populations.

## Supporting information

Supplementary file 1. Interview guide

Supplementary file 2. Interview excerpts

## Data Availability

All data produced in the present study are available upon reasonable request to the authors through DataverseNL.

https://doi.org/10.34894/CZKSKW

## Notes

### Competing Interest Statement

DZ worked as a practice assistant for Jans Huisartsen before commencement of the study.

### Funding Statement

This study did not receive any funding

### Author Declarations

The non-WMO review committee of the Erasmus MC waived ethical approval for this work.

