## Supplementary file 1. Interview guide for "The experiences of general practitioners regarding communication with patients from different cultural backgrounds and/or low socio-economic status: a qualitative interview study"

**Interview guide GP-SES-COM**
**Version 1.0, 30-07-2024**

This interview guide concerns a description of qualitative research through semi-structured interviews for the study: “The experiences of GPs regarding communication with patients from different cultural backgrounds and/or low socio-economic status: a qualitative interview study”.

The duration of the interviews will be about 20-30 minutes.

Participants are approached to ask if they would like to participate in a face-to-face interview following their work as general practitioners at Jans Huisartsen.

**Objective**
The purpose of these interviews is to gain insight into how family physicians experience communication with patients from other cultural backgrounds and/or with low SES, what kind of problems they may experience in doing so, and whether they have suggestions for improvement.

**Preperation**
Discuss before interview starts:
- Introduce interviewer and explain study/purpose of study
- Emphasize that interview is voluntary and participant may stop at any time
- Ask permission to record audio

Turn on audio recording in Teams and start calling date and number of participant.

*“We will process and analyze the responses anonymously so that it cannot be you. Therefore, we will record the interview. This is also done anonymously. In the interview there are no right or wrong answers. You may indicate at any time that you want to stop the interview.”*

General tips for interview:
- Ask through to what is behind the participant's answers. “Can you tell more about that?”, ‘In what respect?’, ‘How does that come about?’, ‘Can you give an example of that?’.
- As much as possible, put yourself in the participant's situation so that the conversation can remain natural.
- Ask open-ended questions
- Do not ask questions in a directing manner
- No own opinion/judgment

**Questions**
1. General information
   a. Age, gender, cultural background
   b. Number of years of experience as a general practitioner, since when working at Jans
   c. In which places worked
2. Experiences communication patients
   a. As a general practitioner, do you interact a lot with patients from different cultural backgrounds and/or low SES?
   b. Can you describe your general experiences in communicating with patients of different cultural backgrounds and/or low SES?
   c. In what areas do you notice differences in communication with patients of the same cultural background and/or low SES?
   d. Are there particular parts of the consultation in which you notice a difference? (history, LO, request for help, shared decision making)
   e. Do you think patients from different cultural backgrounds and/or low SES have different needs?
3. Challenges in communication
   a. What challenges do you encounter when communicating with patients from different cultural backgrounds?
      i. Can you give specific examples or situations in which these challenges became evident?
4. Impact on quality of care

a. How do these communication challenges impact the quality of care you provide?
b. Do you think these challenges impact patient outcomes?
    i. If so, how?

5. Dealing with challenges
    a. How do you deal with the challenges experienced in communicating with these patients?
    b. Do you have particular conversation techniques that you employ with this patient group?
    c. Can you give a specific example in which this strategy worked well?
6. Training in communication
    a. Have you received training/education on communication with patients from different cultural backgrounds and/or with low SES? If so, what was it like?
    b. Do you feel that adequate support and resources are available to help you with these communication challenges?
    c. What additional training or resources would you find helpful?
7. Suggestions for improvement
    a. Based on your experiences, what suggestions do you have for improving communication with patients of different cultural backgrounds and/or low SES?
    b. Are there any policy changes or institutional supports that you think would help address these issues?

8. Closing
    a. Are there any other things in this area you would like to share/disclose?

*"Thank you very much for your cooperation."*
